# A patient-centric characterization of systemic recovery from SARS-CoV-2 infection

**DOI:** 10.1101/2022.06.18.22276437

**Authors:** Hélène Ruffieux, Aimee L. Hanson, Samantha Lodge, Nathan G. Lawler, Luke Whiley, Nicola Gray, Tui Nolan, Laura Bergamaschi, Federica Mescia, Cambridge Institute of Therapeutic Immunology and Infectious Disease-National Institute of Health Research (CITIID-NIHR) COVID BioResource Collaboration, Nathalie Kingston, John R. Bradley, Elaine Holmes, Julien Wist, Jeremy K. Nicholson, Paul A. Lyons, Kenneth G.C. Smith, Sylvia Richardson, Glenn Bantug, Christoph Hess

## Abstract

The biology driving individual patient responses to SARS-CoV-2 infection remains ill understood. Here, we developed a patient-centric framework leveraging detailed longitudinal phenotyping data, covering a year post disease onset, from 215 SARS-CoV-2 infected subjects with differing disease severities. Our analyses revealed distinct “systemic recovery” profiles with specific progression and resolution of the inflammatory, immune, metabolic and clinical responses, over weeks to several months after infection. In particular, we found a strong intra-patient temporal covariation of innate immune cell numbers, kynurenine- and host lipid-metabolites, which suggested candidate immunometabolic pathways putatively influencing restoration of homeostasis, the risk of death and of long COVID. Based on these data, we identified a composite signature predictive of systemic recovery on the patient level, using a joint model on cellular and molecular parameters measured soon after disease onset. New predictions can be generated using the online tool http://shiny.mrc-bsu.cam.ac.uk/apps/covid-systemic-recovery-prediction-app, designed to test our findings prospectively.

**Graphical abstract:** 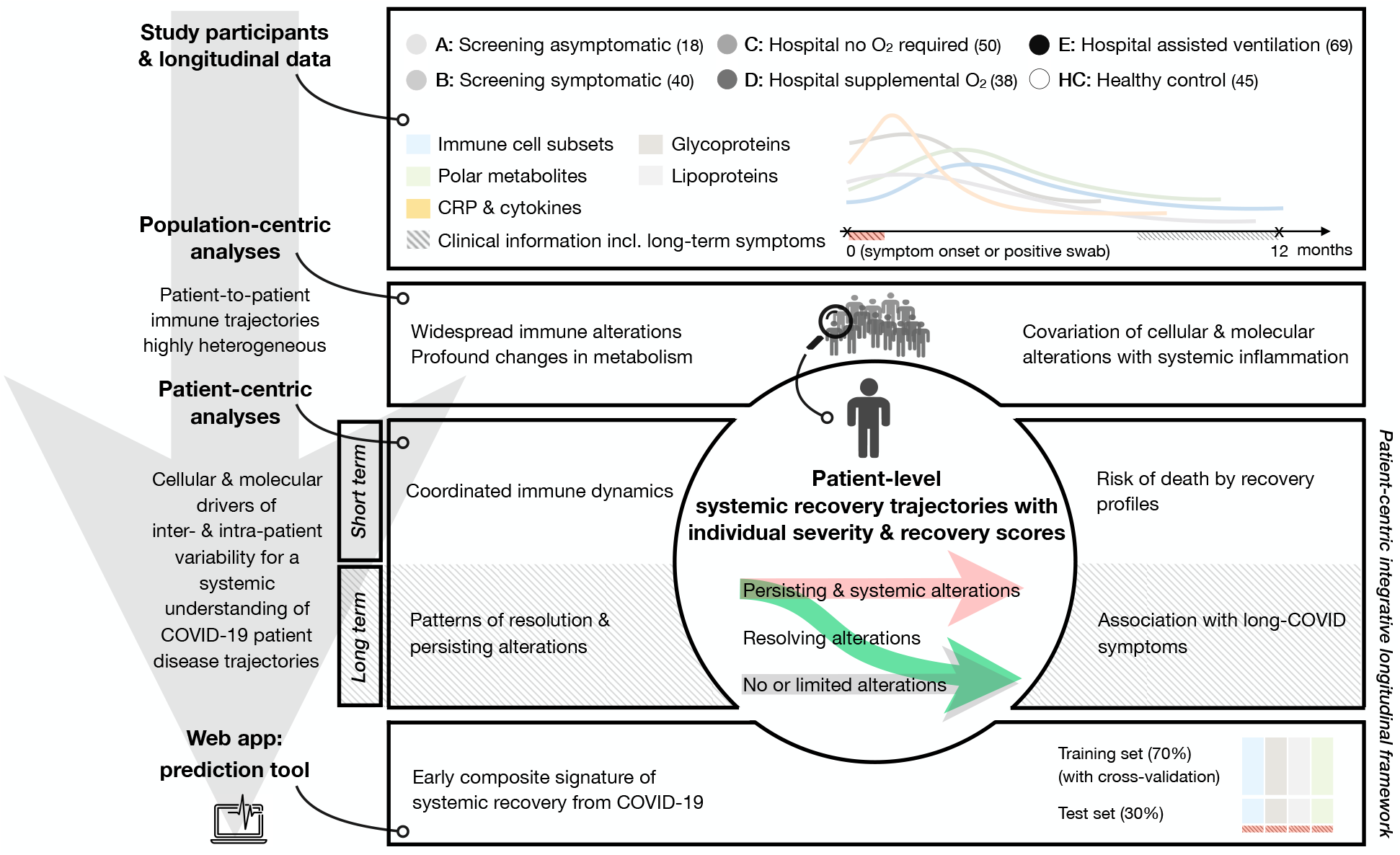

## Introduction

The *coronavirus disease 2019* (COVID-19), caused by the *severe acute respiratory syndrome coronavirus 2* (SARS-CoV-2), has a wide spectrum of clinical manifestations and has led to over 6 million deaths worldwide by mid-May 2022^1^. When acute infection is resolved, health is restored in most individuals, yet some develop prolonged symptoms (long COVID) ^2–4^. Previous work demonstrated that SARS-CoV-2 can induce a significant acute phase reaction (systemic inflammation), profound changes in metabolism, and alterations across many elements of the immune system ^5,6^. Evolution over time of these parameters is highly heterogeneous between patients, with cellular and molecular perturbations persisting for months after the acute phase of the infection and viral clearance in some individuals^7,8^. How failure to restore (immune) homeostasis relates to recovery from acute infection and development of long COVID remains unclear.

Disentangling the interrelation between the individual clinical disease course, and immune cell-, metabolic- and inflammatory alterations is needed to acquire a systemic understanding of COVID-19, both in its acute and chronic form. Such insight may also point at predictors of risk for short and long-term complications of infection, and could help devise strategies for personalized, early intervention.

In this study, we exploited existing and new data from a previously described COVID-19 cohort ^9^, for which immunophenotypes, molecular measurements (including inflammatory markers, polar metabolites, glycoproteins and lipoproteins) and patient questionnaires addressing long-term symptoms of disease have been collected over an extended follow-up period of 12 months. We devised a statistical framework integrating patients’ longitudinal profiles using a two-stage approach. We first evaluated how classical descriptive analyses on the overall disruption of available biological parameters at the population level supported the findings from the initial study and published studies based on independent cohorts. We then estimated the trajectories of the disrupted parameters at the patient level, using longitudinal joint models to borrow strength across data types and patient profiles, and assess coordinated changes over time. Specifically, we deployed a functional principal component (FPC) analysis that examined the cellular, metabolic and inflammatory drivers of inter- and intra-patient variability. We also employed supervised and unsupervised mixed modeling approaches to evaluate parameter recovery up to one year post symptom onset, and probe how they related with survival and self-reported long-term symptoms. Finally, we tested whether a composite signature predictive of systemic recovery could be identified, that would permit risk stratifying new patients. The ambition of this work was to provide actionable insight on individual disease courses to guide future development aimed at improving clinical decision making.

## Results

### Patient data and study design

The study recruited 215 SARS-CoV-2 positive individuals, who were categorized according to five severity classes, A to E, ranging from asymptomatic to severe infection based on their hospitalization status and oxygenation supplementation (Methods). All patients had blood samples taken at study entry and at regular intervals up to one year from symptom onset (classes B to E) or positive swab (class A), permitting detailed longitudinal phenotyping. Comprehensive assays were set up, measuring immune cell subsets, polar metabolites, glycoproteins, lipoproteins, serum cytokines and CRP levels. Follow-up questionnaires assessing long-term symptoms were also obtained from symptomatic patients. The cohort further comprised 45 SARS-CoV-2 negative healthcare workers, to serve as controls and for whom blood for analyses was obtained at a single timepoint.

### Population-level analysis confirms widespread immune and metabolic alterations in association with systemic inflammation

To relate the immunometabolic changes in our cohort to findings from the published literature, we first examined the impact of SARS-CoV-2 infection across clinical, metabolic and cellular variables over a 7-week window post positive swab or symptom onset, using simple linear mixed models accounting for repeated subject measurements (Methods).

A differential abundance analysis between healthy controls and COVID-19 patients across all severity groups indicated widespread systemic dysregulation (Fig. S1), in line with the initial data on our cohort ^9^ and with other reports on independent cohorts ^5,6^. At the population level, there was a metabolic signature in infected patients, where metabolic intermediates from the kynurenine pathway (3-hydroxykynurenine, kynurenine, quinolinic acid) were elevated, while the upstream amino acid, tryptophan, was depleted. Likewise, the abundance of several other amino acids was reduced in plasma from infected subjects. There was also a prominent decrease in high-density lipoproteins (in particular, apolipoproteins), and an increase in very low-density lipoproteins (in particular, triglycerides and free cholesterol) and in the N-acetyl glycoprotein signals GlycA and GlycB. Lastly, as also shown in our previous work ^9^, absolute numbers of neutrophils, plasmablasts and activated CD8^+^ T cells were increased in COVID-19 patients, whilst a marked decrease in some B cell subsets, and classical and non-classical T cell subsets, was also observed in infected individuals. Association analyses with C-reactive protein (CRP) serum concentration — an indicator of the acute phase response in COVID-19 patients ^10,11^ — further indicated significant linear relationships with most metabolic and cellular variables (Fig. S2).

### Disease severity and recovery profiles account for most of the inter-patient variation in the systemic inflammatory response

The above analyses found marked molecular and cellular abnormalities across COVID-19 patients and extensive covariation between systemic inflammation and disrupted parameters. Population-level exploration fails, however, to provide patient-level longitudinal insight. To overcome this limitation, we applied a functional principal component (FPC) framework to the parameter signatures discussed above to (1) disentangle the molecular heterogeneity in patients’ response to SARS-CoV-2, and (2) provide patient-specific longitudinal estimates of the extent and dynamics of recovery, along with a characterization of the main factors associated with the disease course.

First, we aimed to identify the main modes of variation in trajectories of CRP (i.e., systemic inflammation) among symptomatic patients (i.e., severity classes B to E) over the first 7 weeks post symptom onset (Methods). The first two FPC eigenfunctions jointly accounted for more than 99% of the variance and, along with their corresponding scores, they were interpretable in terms of the patients’ disease profiles (Fig. 1A-B). Namely, the first eigenfunction acted as a proxy for inflammation severity, with associated *“severity” patient scores* (*x*-axis, Fig. 1B): patients with positive severity scores had higher than average CRP levels over the 7-week period (e.g., patient CV0047 in Fig. 1B), whilst the opposite held true for patients with negative scores (e.g., patient CV0046 in Fig. 1B). This interpretation was independently corroborated by the B-to-E severity classes assigned to each patient based on their hospitalization and oxygenation support. The second eigenfunction acted as a proxy for recovery from inflammation, with associated *“recovery” patient scores* (*y*-axis, Fig. 1B): patients with large positive scores for the second eigenfunction had a drastic improvement of their inflammatory status over time (e.g., patient CV0115 in Fig. 1B), whilst inflammation deteriorated or resolved more slowly compared to the population mean function in patients with negative scores (e.g., CV0212 in Fig. 1B). Hierarchical clustering of the severity and recovery scores uncovered three patient groups with distinct inflammation trajectories (Fig. 1B-C): absent or mild inflammation over the 7-week window post symptom onset (group *i*), early, resolving inflammation (group *ii*), and persisting inflammation (group *iii*). This confirmed that the patient FPC estimates encompass information on the magnitude and temporal profile of the systemic inflammatory infection response as captured by CRP measurements. Sex and age distribution in each group aligned with existing knowledge on COVID-19 risk factors ^12^ (Fig. 1D-E). Notably, the groups were not entirely driven by disease severity, but also by the type of recovery profile. In particular, patients in group *ii* had higher than average *y*-axis recovery scores, and each of the three groups was comprised of patients from multiple severity classes (Fig. 1B). We thus refer to this new patient classification as *recovery groups*.

**Figure 1:**
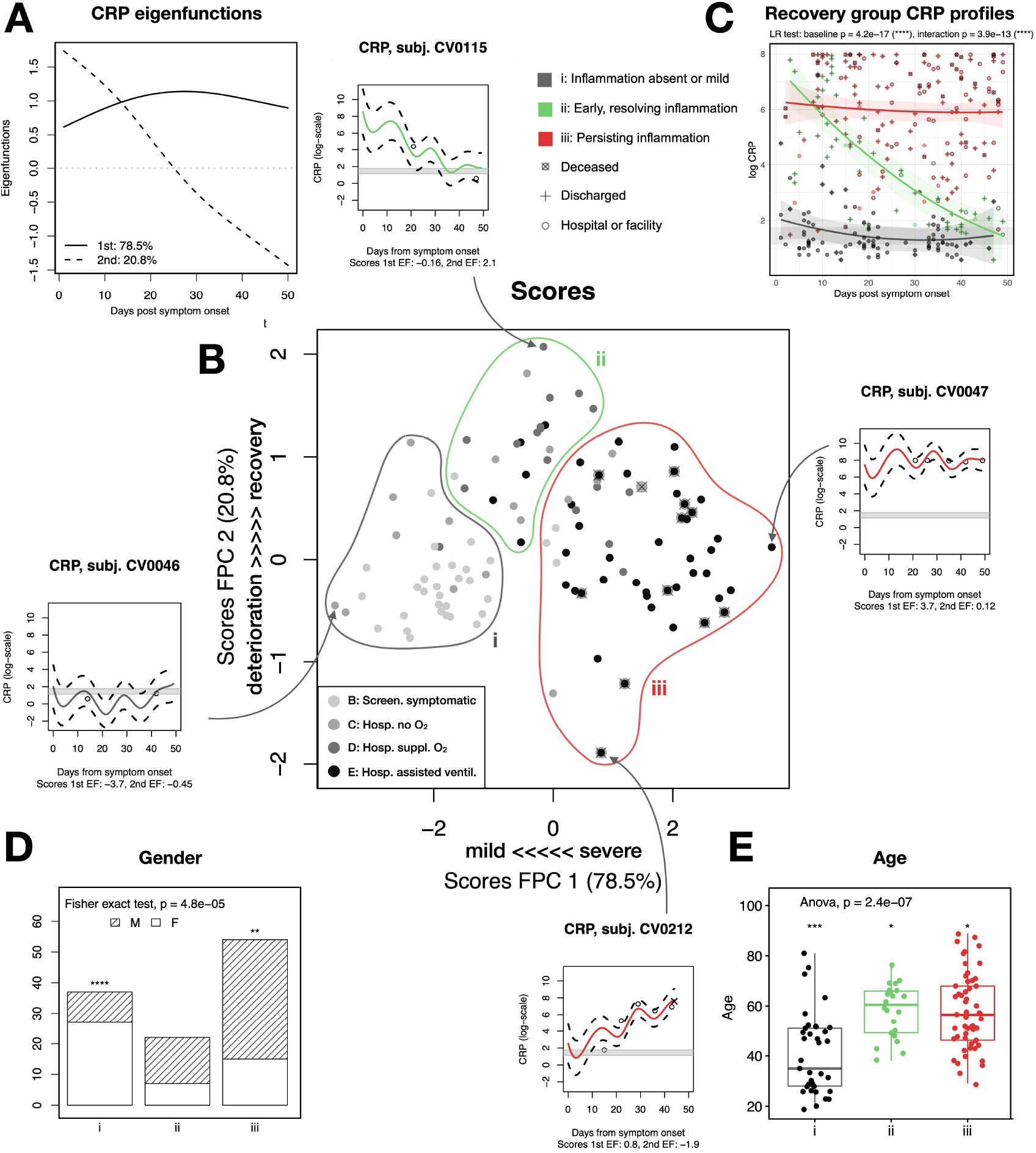
Functional principal component (FPC) analysis of COVID-19 patients’ CRP levels. A: First two eigenfunctions representing the severity of inflammation and the recovery from inflammation, respectively, over the 7-week window of the FPC analysis, and accounting for 78.5%, respectively 20.8% of the variability. B: Scatterplot showing the scores corresponding to the first and second eigenfunctions; each point corresponds to one patient. Severity scores are on the *x*-axis (the higher the more severe the inflammation) and recovery scores are on the *y*-axis (the higher the more pronounced the temporal resolution of inflammation). The legend B-to-E indicates the study severity class assigned to each patient based on their hospitalization status and oxygenation supplementation; the grey-color gradient observed on the *x*-axis suggests that these classes are reflected by the estimated severity scores. The lines delineate the three “recovery groups” *i, ii* and *iii*, obtained by hierarchical clustering. The snippets show log-transformed CRP trajectories for four examples of patients with extreme severity or recovery scores. The grey bands indicate normal CRP levels: they correspond to the interquartile range of healthy controls’ (log-transformed) CRP levels. The points correspond to the observed values, the red, green and grey curves are the trajectories estimated using the FPC model and the dashed curves delineate the 95% confidence bands. C: CRP trajectories conditional on the three recovery groups with 95% confidence bands, estimated with a longitudinal mixed model accounting for patients’ repeated measurements (likelihood ratio tests for the baseline group effect and group *×* time interaction effect). Points correspond to observed values. D: Characterization of the recovery groups by gender (one vs all and overall Fisher exact test). E: Characterization of the recovery groups by age (one vs all *t*-tests and anova).

### Joint analysis of molecular parameters sheds light on their coordinated dynamics during acute infection and convalescence

The strong association between circulating CRP concentration and components of the immune response to COVID-19 has been extensively characterized (see, e.g., our previous work ^9^ or the above population-level analyses), as has the link between inflammation and clinical severity ^8,10,11^. By contrast, how longitudinal post-infection profiles — e.g., as reflected by groups *i, ii* and *iii* (Fig. 1C) — relate with organismal recovery has not been previously explored. To start addressing this, we first examined the immune and metabolic profiles of groups *i, ii* and *iii* by analyzing the series of available parameters longitudinally over the 7-week window following symptom onset. Mixed-effect modeling with time encoded as a continuous variable revealed that the trajectories of many cellular and molecular parameters largely reflected the inflammation profile that characterized each recovery group (examples shown in Fig. 2A). For instance, quinolinic acid levels remained mostly unperturbed for group *i*, they were increased early but returned to baseline levels at later timepoints for group *ii*, and they started and remained high weeks after symptom onset for group *iii*. To explore the interplay between the temporal profiles of the inflammatory, immune-cell and metabolomic responses on the patient level, we performed additional multivariate FPC analyses on sets of parameters whose alterations were found to be signatures of active SARS-CoV-2 infection in our above population-level analyses, namely, serum cytokines, lymphocyte subsets, apolipoproteins, glycoproteins and kynurenine-pathway metabolites (Methods). Trajectories reconstructed for each patient largely co-evolved over the disease course, as illustrated on a selection of parameters for three patients from group *i, ii* and *iii* (Fig. 2B). The quinolinic acid temporal profiles for these patients broadly agreed with the group-level longitudinal estimates discussed above (Fig. 2A). More generally, the 95% confidence bands of the estimated trajectories for group-*i* patient CV0261 (symptomatic but not hospitalized) covered the normal levels (healthy-control interquartile-range grey bands), suggesting absent or mild alterations. The trajectories of group-*ii* patient CV0115 (with supplemental oxygen) were slightly above or below normal levels, yet with no sign of deterioration over time, but hints of normalization, in particular for plasmablast levels. Finally, group-*iii* patient CV0212 (with assisted ventilation and who died 44 days post symptom onset) exhibited a clear deterioration in all parameters, with the estimated trajectories departing from normal levels. Although the parameter courses of these three patients tended to be representative of the general parameter evolution within the groups, some patients displayed peculiar parameter trajectories, again emphasizing the value of patient-specific estimates to resolve the covariation of cellular/molecular parameters with inflammation at the individual level. The reconstructed cellular, molecular and inflammation trajectories for all patients, and the group-level trajectories can be inspected online at http://shiny.mrc-bsu.cam.ac.uk/apps/covid-patient-trajectories.

**Figure 2:**
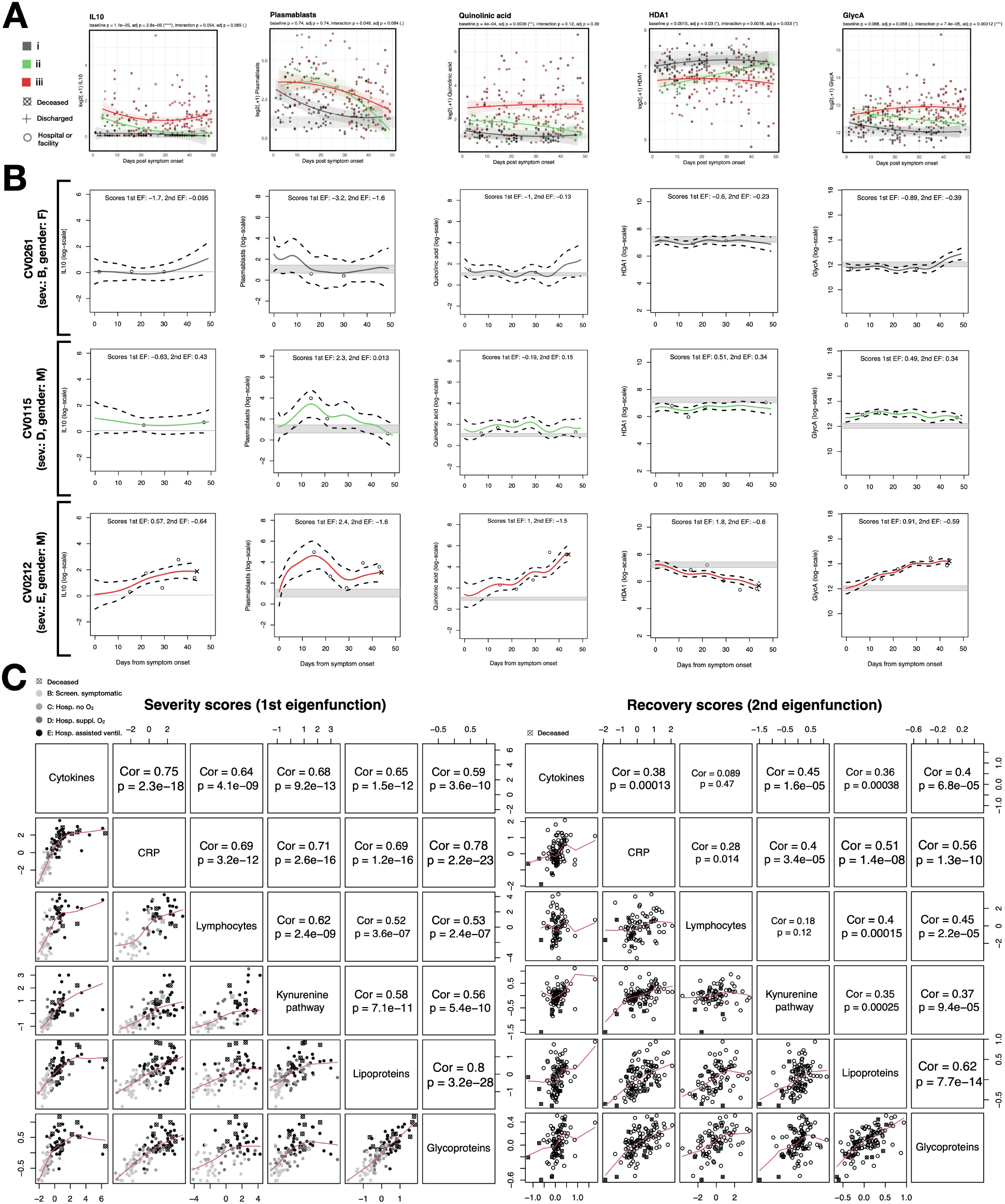
Group-level and patient-level estimates of cellular and molecular trajectories: cytokines, CRP, lymphocytes, lipoproteins, glycoproteins and kynurenine-pathway metabolites over the first 7 weeks post symptom onset. A: Recovery-group trajectories estimated by longitudinal mixed modeling for five parameters selected from each data type. All levels have been log-transformed and the grey bands correspond to the interquartile range of healthy controls’ levels. Adjusted *p*-values from likelihood ratio tests for baseline and interaction effects are indicated. B: Trajectories of the same parameters estimated by FPC analysis for three COVID-19 patients, from each recovery group. C: Comparison of the severity and recovery scores obtained from the six FPC analyses with Pearson correlation test.

To further explore the covariation of parameters over time across data types in each recovery group, we conducted correlation analyses on all variables analyzed by FPC (Methods), stratifying the patient data into (1) acute-infection phase (less than 3 weeks post symptom onset) and (2) protracted-infection/convalescence phase (3 *−* 7 weeks post symptom onset). The correlation patterns for all three patient groups in both time bins tended to be stronger than those of healthy controls (Fig. S3). This suggested that, irrespective of disease severity, as a population, infected individuals did not make full organismal immune and metabolic recovery at late phases of the disease. The magnitudes of the correlation between molecular and cellular components again reflected the inflammation profile characterizing each recovery group (Fig. 1C). A detailed inspection of specific pairwise patterns revealed that, for group *iii*, kynurenine-pathway metabolites and cytokines (i.e., systemic inflammation) were positively correlated, whereas such covariation was not observed in recovery groups *i* and *ii*. Aligning with the protracted lymphopenia typically observed in patients with persistent inflammation, there were also significant positive pairwise correlation patterns among lymphocytes (CD4 EMRA, CD4 Naive, CD4 Non-naive HLA-DR^+^CD38^+^ T, CD8 EMRA, CD8 Naive, CD8 Non-naive HLA-DR^+^CD38^+^ T, CD19^+^, plasmablasts, gd T, MAIT, NK & NKT cells) in both time windows.

On top of bringing parameter-specific insights, our additional FPC analyses more broadly defined the interplay between data types in response to infection. Similarly to the CRP FPC analysis, the first and second eigenfunctions could be interpreted as proxies for “severity” and “recovery” (or “normalization”) of the molecular and cellular trajectories, respectively (Fig. 2C). The different sets of severity scores were highly correlated, confirming that the inflammatory, immunologic and metabolomic alterations were closely interlinked. Moreover, these scores again largely echoed the clinical severity classes B-to-E, indicating that the variability in the various trajectories reflected pathophysiologic signatures relevant to clinical disease. The correlation of recovery scores across the different types of parameters was somewhat weaker, although still significant in most cases. This may reflect distinct dynamics between cellular compartments or differing half-lives. Alternatively, this could indicate persistent disruption of biologic systems despite resolution of inflammation, a hypothesis we explored below. Nevertheless, the overall covariation across FPC scores provided a strong rationale to (1) model all cellular and molecular data in a joint manner, in order to borrow strength across parameters that co-evolve over time as regulated by shared biological processes, and (2) train integrative models exploiting cellular and molecular parameters to predict organismal recovery at the individual patient level.

### Long-term molecular characterization of recovery profiles: patterns of resolution and persistent alterations

The above analyses highlighted important variability across cellular and molecular patient trajectories over the first 7 weeks post symptom onset. Using mixed modeling, we next examined the long-term dynamics of cellular and molecular recovery over one year after infection (Methods).

As expected, group *i* (with mild or absent inflammation) continued to show minor or absent disruptions over this extended follow-up period (Fig. 3). There were, however, exceptions to this, with significant differences beyond 12 weeks post symptom onset for some metabolites, compared with healthy controls, likely driven by few individuals with persisting abnormalities (possibly associated with long-term clinical manifestations as discussed below). In group *ii*, many parameters were altered early on, i.e., up to 3 weeks post symptom onset for most, and up to 7 weeks for some (including several lipoproteins, glycoproteins and amino acids), but were later indistinguishable from healthy controls — thus aligning with the recovery from inflammation defining the group. Conversely, group *iii* (persistent inflammation) showed widespread and long-lasting cellular and molecular alterations. These observations linked, on the population level, systemic inflammation and immunometabolic dysregulation up to one year post infection. We also assessed the baseline (i.e., close to symptom onset) group effect, and the group *×* time interaction effect, using the group-level longitudinal mixed models discussed above (Fig. 2A). For a number of parameters, significant baseline and/or interaction effect(s) were detected. As tested below, these parameters, when measured early post infection, might contain information regarding a patient’s ability to recover.

**Figure 3:**
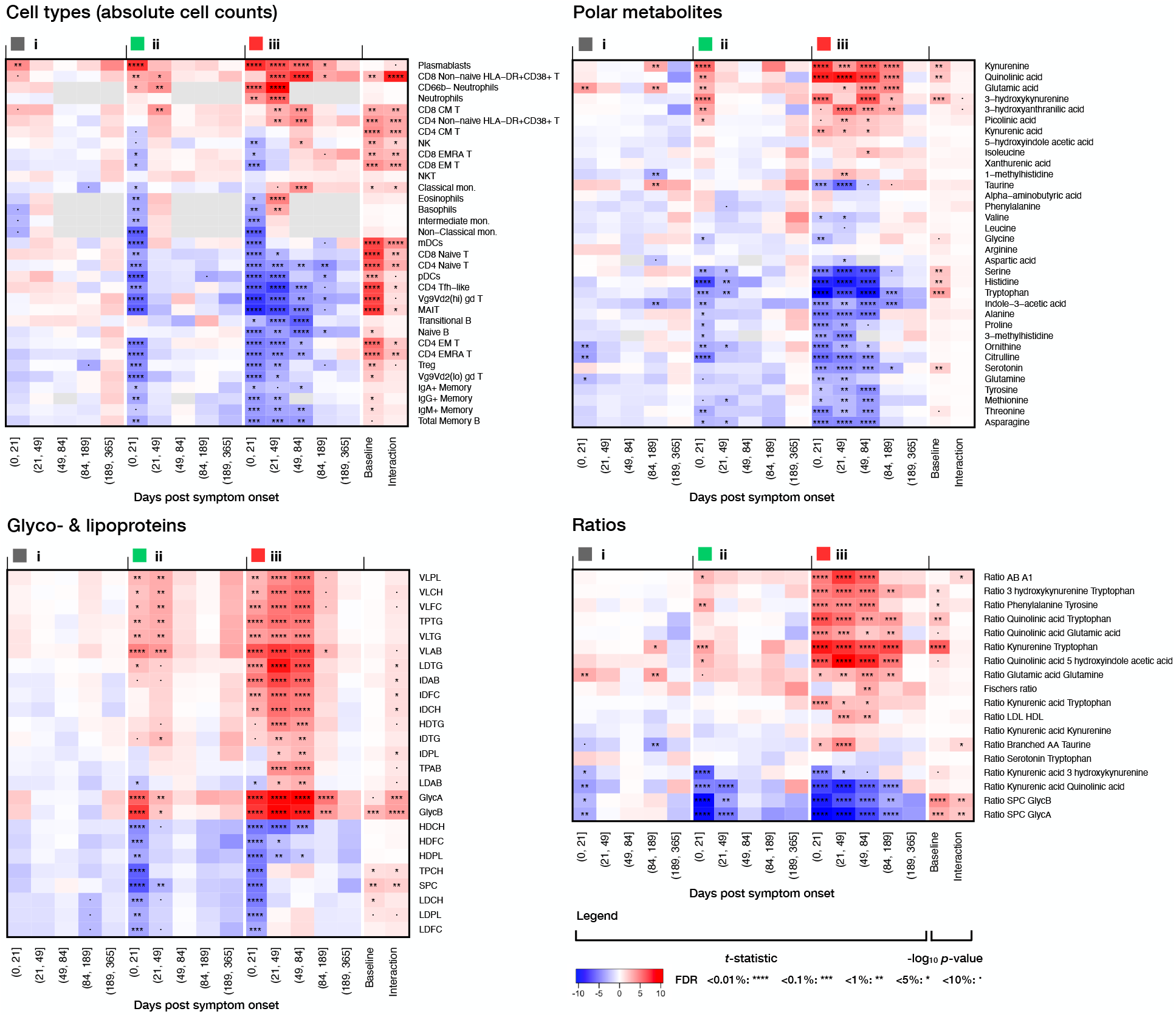
Long-term recovery-group trajectories for immune cell subsets, polar metabolites, main classes of glyco-& lipoproteins and diverse metabolic ratios. Comparison of the parameters of each recovery group with the healthy control levels over a year post symptom onset, using mixed modeling. The first 5 *×* 3 columns indicate the *t*-statistics obtained for the group effect and corresponding significance after FDR multiplicity adjustment, and the last two columns indicate the significance (*−* log_10_ adj. *p*-value) of the baseline and interaction effects using the group-level longitudinal mixed models.

### Clinical associations with recovery profiles: long-COVID symptoms and risk of death

We next asked how the recovery profiles related to long COVID, using questionnaires on long-term symptoms collected from patients between 2 and 11 months post symptom onset (Methods). A comparison of the reported symptoms demonstrated significant differences across recovery groups (Fig. 4A). In particular, patients in group *iii* reported more neurological symptoms (fatigue, muscle weakness, pain, difficulty eating, drinking, swallowing) compared to group *i*. In group *iii*, fatality implied that the reported symptoms only reflected the subpopulation of patients who were alive several weeks after infection, and mechanical ventilation was a source of non-infection related sequelae, which added (unavoidable) limitations. We also computed composite scores by averaging the different severity scores, and compared these scores across recovery groups. Again, patients from group *iii* had a significantly poorer composite score compared to the other two groups, while the converse was true for patients from group *i*. There was, however, substantial variability within each recovery group, in particular, a few patients in groups *i* and *ii* had high composite scores, although their cellular, inflammatory and molecular trajectories — as estimated by our FPC analyses — had essentially returned to normal levels within 7 weeks post symptom onset. This suggested persistent systemic subjectively perceived sequelae for these subjects despite absent, or rapidly resolving inflammation and cellular/molecular disruption.

**Figure 4:**
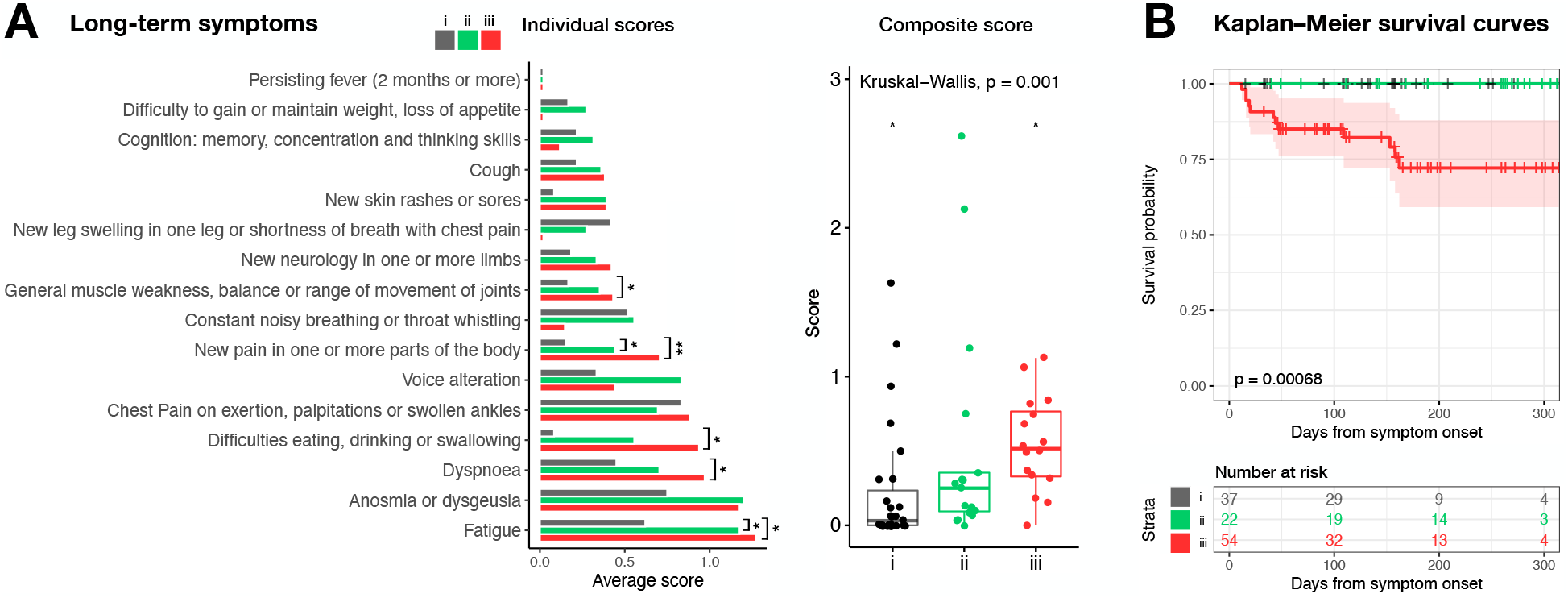
Long-term symptom characterization of the recovery groups and survival probabilities. A: Average individual scores for long-term symptoms with pairwise Wilcoxon test, and symptom composite scores per recovery group with one vs all Wilcoxon tests (stars) and overall Kruskal—Wallis test. B: Kaplan—Meier survival curves and table for the three recovery groups, with log-rank test *p*-value. The curves for groups *i* and *ii* overlap (green and black).

Assessing how recovery profiles related to mortality showed that all 12 patients who died belonged to group *iii* (Fig. 1B). A survival analysis confirmed the significance of the increased risk of death for this group compared to groups *i* and *ii*, when accounting for the number of observations and drop-out events (Fig. 4B).

### Clinically testable predictive signatures of systemic recovery

Our FPC analyses identified the inflammation recovery groups *i, ii* and *iii* as three categories of disease trajectory. Follow-up analyses further indicated that these groups reflected *systemic recovery* from COVID-19, by showing that the patients’ inflammatory response to infection was tightly linked with long-term clinical consequences and with the magnitude and temporal resolution of immunometabolic abnormalities, over months post symptom onset. We next aimed to test whether this insight could be leveraged to predict, soon after infection, the recovery profile of individual patients, by training an integrative model on samples collected during the early phase of the disease. We applied a generalized canonical correlation algorithm, extended for supervised analysis, on all cellular and molecular data jointly (Methods). The method identified composite signatures of systemic recovery via an internal selection of parameters relevant for prediction. Specifically, it implemented a trade-off between (1) maximizing the correlation of biological parameters (immune cell subsets, polar metabolites, glycoproteins, lipoproteins and diverse metabolic ratios) and (2) maximizing the discrimination between unfavorable recovery profiles (group *iii*) and favorable recovery profiles (groups *i & ii* merged). We used the first sample of each patient, provided it was taken within 3 weeks from symptom onset, and relied on training-test set splits involving 70% and 30% of the samples, respectively. The signatures for the first two latent components (Fig. 5A) and the circular plot (Fig. 5B) were obtained using the training samples, while the receiver operating characteristic (ROC) curves (Fig. 5C) were obtained using the left-out test samples.

**Figure 5:**
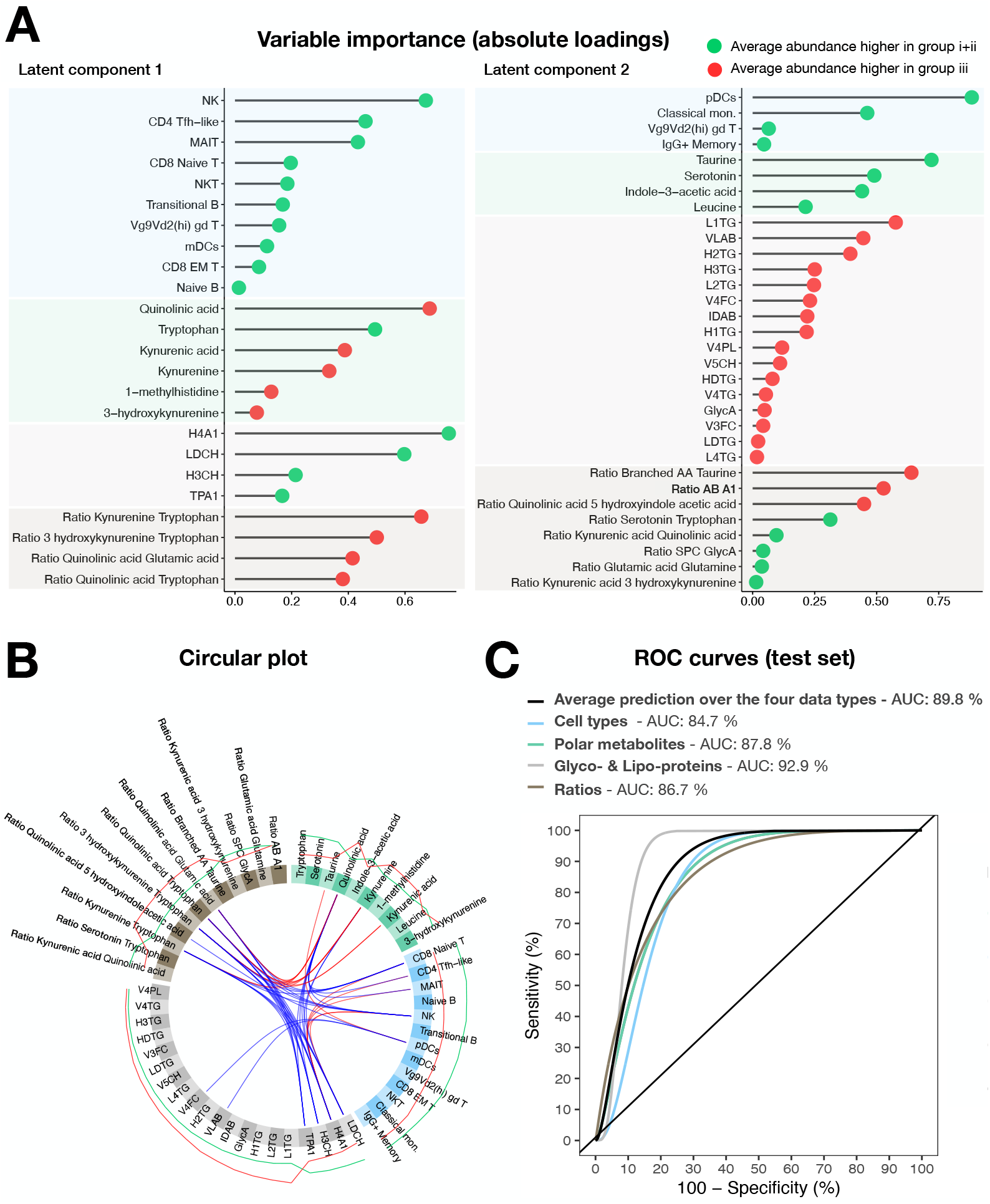
Predictive modeling for the recovery groups (*i+ii* and *iii*). A: Absolute loadings forming early predictive signatures, for the first and second latent components of the sparse generalized canonical correlation analysis (sGCCA) model, arranged per data types. The green and red dots indicate that the average abundance is greater in group *i+ii* or group *iii*, respectively. B: Circular plot linking pairs of variables from the two signatures, if their absolute Pearson correlation exceeds 0.75, with red and blue indicating a positive and negative correlation, respectively. The external green and red lines show the relative average abundance of the selected variables within the two categories (green: groups *i+ii* and red: group *iii*). C: ROC curves for the predictive performance in classifying individuals from the test set into group *i+ii* or group *iii*, using the sGCCA model. The curves show the average prediction based on the four data types (black), and the prediction based each data type separately (colors).

The sign of the average signature parameter abundances aligned with existing knowledge and with the analyses presented in Fig. 3. Metabolic intermediates from the kynurenine pathway, and corresponding ratios, again appeared as important markers of the type of recovery, corroborating previous findings on the involvement of the pathway in immunity and inflammation^13^. Quinolinic acid, tryptophan, kynurenic acid, kynurenine and 3-hydroxykynurenine were selected in the first signature, while serotonin, a neurotransmitter derived from tryptophan catabolism, was selected in the second signature. These metabolites appeared together with a series of innate immune cells.

Lastly, the presence of high-density lipoproteins in the first signature and the large triglyceride contribution in the second signature echoed the results obtained above (Fig. 3). The area under the curve (AUC) on the test set reached 89.8% (Fig. 5C). The predictive performance was also good when restricting the signatures to each data type (AUCs *>* 84.7%), further suggesting high interdependence of the biologic processes captured in our study.

Finally, we developed an interactive tool to browse predictions based on selected markers from the two systemic recovery signatures: http://shiny.mrc-bsu.cam.ac.uk/apps/covid-systemic-recovery-prediction-app (Methods). Each new prediction obtained for a patient presented to the clinic also entails an estimated prediction score to convey the level of confidence in the predicted recovery profile, and the predictive performance on the test set is reevaluated based on the subset of markers supplied. This pilot tool aims to facilitate the planning of prospective studies for testing the generalizability and clinical actionability of our findings.

## Discussion

COVID-19 is a heterogeneous disease with strong patient-to-patient variability of the immune, metabolic and inflammatory response over time. A growing literature reports widespread cellular and molecular abnormalities in association with systemic inflammation in different study cohorts, yet population-level analyses are unable to resolve individual patient disease courses. Here, we deployed a patient-level framework on longitudinal immunophenotyping, metabolic and clinical data from patients with differing disease severity to study recovery from COVID-19 in a broad, systemic sense.

### Estimating the cellular and molecular dynamics and their interplay at the patient level sheds light on recovery profiles

Our framework relied on a multi-data-type FPC approach, which refined conventional population-level analyses in several respects. First, it allowed reconstructing the inflammatory, metabolic or immune trajectories of each patient during acute infection and convalescence using multivariate modeling, thereby borrowing strength across parameter temporal coregulation patterns. Second, it estimated scores that captured the inter-patient parameter variability over time, in relation with the individual severity and recovery profiles; this estimation was completely data-driven, i.e., it made no use of predefined severity classes or any other *ad-hoc* information. Third, a clustering analysis of the CRP FPC scores uncovered three types of patient profiles that were distinctive not only in the dynamics of inflammation, but also across a range of cellular and metabolic parameters over months post symptom onset, as well as in terms of risk of death and long-term symptoms. These profiles therefore constituted distinct patient systemic recovery categories, that characterized disease courses beyond clinical severity (for which molecular correlates and risk factors have already been extensively described by us and others ^5,9,11,14^). The patient trajectories, scores and correlation functions can be visualized interactively (http://shiny.mrc-bsu.cam.ac.uk/apps/covid-patient-trajectories).

### Systemic inflammation, perturbed kynurenine-pathway and innate immune activity, and incomplete recovery

The estimated temporal covariation patterns across data types suggested that acute-phase inflammation is a common denominator interlinked with incomplete clinical and immunometabolic recovery, up to a year post disease onset. While this observation unsurprisingly adds COVID-19 to the long list of diseases in which inflammation plays a central role^15^, our analyses highlighted that the coordinated dynamics of the innate immune system, kynurenine and host lipid metabolism are likely pivotal in restoring overall homeostasis. In particular, our data suggested that a limited number of pathophysiological processes impact many of the parameters appearing together in the composite predictive signatures (http://shiny.mrc-bsu.cam.ac.uk/apps/covid-systemic-recovery-prediction-app). For instance, natural killer (NK) cells, with the largest weight in the first signature, play a central role in anti-viral immunity through the secretion of pro-inflammatory cytokines and cytotoxic activity ^16^. The clear reduction of NK cells in peripheral blood from subjects with unresolving CRP (group *iii*) suggests an inflammation-driven perturbation of the NK cell compartment. The kynurenine pathway is also intricately associated with inflammation, as it is activated through indoleamine 2,3-dioxygenase (IDO) induction by pro-inflammatory cytokines. It has also been suggested as a key pathway involved in mechanisms linking inflammation and central nervous system alterations, by favoring the degradation of tryptophan towards 3-hydroxykynurenine and quinolinic acid (both appearing in the first predictive signature), and by reducing serotonin production (second signature). Tryptophan catabolism into neuroactive kynurenine metabolites and serotonin highlights its importance in regulation of neural function. Kynurenine can be metabolized into 3-hydroxykynurenine and further processed into quinolinic acid (QA). QA is an NMDA receptor agonist that exhibits similar potency as endogenous agonists, glutamate and aspartate. Binding of QA to NMDA receptors results in substantial Ca2^+^ flux into neurons, which characteristically results in cell death. By contrast serotonin or 5-hydroxytryptamine (5-HT) is a neurotransmitter derived from tryptophan catabolism involving tryptophan hydroxylation and decarboxylation. In patients with unresolved inflammation, the marked reduction in serotonin over the course of the disease suggests that tryptophan degradation is skewed towards the kynurenine pathway. Hence, it is plausible that abnormal levels of kynurenine pathway intermediates coupled with the significant reduction of serotonin abundance may contribute to the neurologic sequelae (e.g., fatigue, weakness, chronic pain) of long COVID, as reported by patients in the poorest recovery group *iii*.

### Early, individual-patient prediction of unfavorable disease course and of long COVID

Whilst mechanistic insight will require additional dedicated research, an important finding of our work is that kynurenine-pathway metabolites measured early post symptom onset tend to have, alone, very good predictive value for the type of systemic recovery profile. In particular, high levels of quinolinic acid, kynurenic acid, kynurenine, 3-hydroxykynurenine, low levels of tryptophan (first signature) and low levels of serotonin (second signature) appeared to be early markers of poor prognosis, hence following the same lines as known associations with clinical severity. Moreover, the predictive performance was further increased by a simultaneous inspection of cellular, metabolic and glyco-& lipoproteomic parameters from the composite signatures. Of note, however, our internal performance assessment on a left-out test set also indicated that minimal subsets from these signatures (involving 3 *−* 4 parameters) may achieve sufficient predictive accuracy to distinguish favorable from unfavorable disease trajectories. These findings also suggested that complementing routine blood test panels with a limited set of parameters, identified here to be of relevance for predicting disease progression, may be of clinical value. We stress however that our prediction framework should not be used as a diagnostic tool, but rather as a pilot study to guide future implementation. The excellent performance in our cohort warrants independent validation; it suggests that such predictive modeling, solely based on early cellular and molecular measurements, could effectively estimate the probability of becoming severely ill or of developing long COVID, and ultimately help support, e.g., decisions on whether to administer antiviral drugs to patients with risk factors, *etc*.

### A basis for monitoring changes in new patient recovery profiles — immunity status and new variants

Our cohort involved unvaccinated patients infected by the Wuhan wild type variant and therefore constituted a clean floor for studying the immunometabolic response triggered by the original strain of the SARS-CoV-2 virus. As new variants emerge, however, and immune systems of individuals are influenced by their own history of vaccination and disease, integrative longitudinal studies such as ours should be repeated to evaluate the stability of the identified cellular and molecular signatures in this new context, monitor the changes in specific biomarkers and in patient profiles, and understand the drivers of these changes. Our patient-level methodology is transferable to any new cohort, which permits such systematic comparative work.

Altogether, our framework constitutes a setup for studying the molecular and clinical correlates of organismal recovery. It offers patient-centric lenses to dissect the heterogeneity of the immune and metabolic responses to SARS-CoV-2 infection, formulate mechanistic hypotheses on their orchestrated action in driving systemic recovery, and develop personalized early intervention strategies.

## Methods

### Cohort and samples

The cohort and initial sampling timeline have been previously described in earlier work ^9^. Late-timepoint samples have subsequently been collected and new metabolic data have been quantified. Briefly, study participants were patients attending Addenbrooke’s Hospital, Royal Papworth Hospital NHS Foundation Trust or Cambridge and Peterborough Foundation Trust with a confirmed diagnosis of SARS-CoV-2 infection, as well as SARS-CoV-2 positive health care workers recruited from a staff screening program. The initial sampling program collected blood samples for 201 patients at study entry (first samples collected 31/3/2020) and at regular intervals up to three months post symptom onset. Late-timepoint samples were then obtained up to a year post symptom onset, approximately at 3, 6 and 12 months following recruitment. For this long-term follow-up, 14 additional hospitalized patients were included alongside the original cohort, after their discharge from Addenbrooke’s Hospital. Each participant was assigned to one of following categories of clinical severity: (A) asymptomatic healthy workers (18 individuals); (B) symptomatic healthy workers (still working or self-isolating, 40 individuals); (C) patients who presented to hospital but never required oxygen supplementation (50 individuals); (D) patients who were admitted to hospital and whose maximal respiratory support was supplemental oxygen (38 individuals); and (E) patients who at some point required assisted ventilation (69 individuals). Controls (45 individuals) were SARS-CoV-2 negative hospital staff members with a negative serology. Blood samples were drawn in EDTA, sodium citrate, serum and PAXgene Blood RNA tubes (BD Biosciences) and processed by the CITIID-NIHR COVID BioResource Collaboration group. All study participants provided written informed consent prior to enrolment. Ethics approval was obtained from the East of England – Cambridge Central Research Ethics Committee (“NIHR BioResource” REC ref 17/EE/0025, and “Genetic variation AND Altered Leucocyte Function in health and disease - GANDALF” REC ref 08/H0308/176).

### CRP and cytokines

High sensitivity C-reactive protein (CRP) and serum cytokines (IL-6, IL-10, IL-1*β*, TNF and IFN-*γ*) were quantified by laboratories in Cambridge using standard assays ^9^.

### Flow immunophenotyping and CyTOF assays

The assays are detailed in previous work on the early follow-up of the study cohort ^9^. Briefly, peripheral blood mononuclear cells (PBMCs) were obtained from peripheral venous blood collected in 10% sodium citrate tubes (up to 27 mL per sample). They were isolated using Leucosep tubes (Greiner Bio-One) with Histopaque 1077 (Sigma) by centrifugation at 800x g for 15 min at room temperature. PBMCs at the interface were collected, rinsed twice with autoMACS running buffer (Miltenyi Biotech) and cryopreserved in FBS with 10% DMSO. All samples were processed within 4 hours of collection.

An aliquot of whole blood (50*µl*) was added to BD TruCount tubes with 20*µl* BD Multitest 6-color TBNK reagent (BD Biosciences) for direct enumeration of T, B and NK cells, and was processed as per the manufacturer’s instructions. Samples were gated in FlowJo v10.2 and the number of cells falling within each gate was recorded. For analysis, these were expressed as an absolute concentration of cells per *µl*, calculated using the proportions of daughter populations present within the parent population determined using the BD TruCount system.

The protocol used to isolate PBMCs led to an impaired recovery of the monocyte populations, specifically intermediate and non-classical monocytes. Measurements were extended to these and other granulocyte populations using a mass cytometric assay for a subgroup of patients and healthy controls (249 samples).

### NMR spectroscopy and mass spectrometry based quantitative metabolic phenotyping

#### ^1^H NMR sample preparation

Sample processing was performed according to Bruker IVDr protocols ^17^ and previously published recommended procedures for IVDr metabolic analysis of COVID-19 plasma samples ^18^. Plasma samples were stored at *−*80^*°*^C until required, after defrosting the samples were centrifuged at 13 000g for 10 min at 4^*°*^C. The plasma supernatant was mixed with buffer (75 mM Na2HPO4, 2 mM NaN3, 4.6 mM sodium trimethylsilyl propionate-[2,2,3,3-2H4] (TSP) in 80% D2O, pH 7.4 *±* 0.1) (1:1). 600*µ*L of the plasma/buffer mixture was transferred into a Bruker SampleJetTM NMR tube (5mm) and sealed with POM balls added to the caps.

#### ^1^H NMR spectroscopy data acquisition and processing parameters

NMR measurements were performed on a Bruker 600 MHz Avance III HD spectrometer (IVDr) equipped with a BBI probe and fitted with Bruker SampleJetTM robot with the cooling system set to 5^*°*^C. A quantitative calibration was completed prior to the analysis ^19^. For each sample a 1H 1D experiment with solvent pre-saturation (32 scans, 98 304 data points, spectral width of 18 028.85 Hz) was completed in automation with an experiment time of 4.5 mins. Lipoprotein reports itemising 112 lipoprotein parameters for each plasma sample were generated using the Bruker IVDr Lipoprotein Subclass Analysis (B.I.LISATM) method. This was completed by quantifying the -CH2 (*δ* = 1.25) and -CH3 (*δ* = 0.8) peaks of the 1D spectrum after normalization to the Bruker QuantRefTM manager within TopspinTM using a PLS-2 regression model. The lipoprotein subclasses included different molecular components of very low-density lipoprotein (VLDL, 0.950–1.006 kg/L), low-density lipoprotein (LDL, density 1.09–1.63 kg/L), intermediate-density lipoprotein (IDL, density 1.006–1.019 kg/L), and high-density lipoprotein (HDL, density 1.063–1.210 kg/L). The LDL subfraction was further divided into 6 density classes (LDL-1 1.019–1.031 kg/L, LDL-2 1.031–1.034 kg/L, LDL-3 1.034–1.037 kg/L, LDL-4 1.037–1.040 kg/L, LDL-5 1.040–1.044 kg/L, LDL-6 1.044–1.063 kg/L), and the HDL subfractions placed in 4 different density classes (HDL-1 1.063–1.100 kg/L, HDL-2 1.100–1.125 kg/L, HDL-3 1.125–1.175 kg/L, and HDL-4 1.175–1.210 kg/L). Upon completion of the standard 1D experiment, the DIRE experiment was run (64 scans, 98 304 data points, spectral width of 18 028.85 Hz) with a total experiment time 4 minutes 25 seconds per sample ^20^.

#### NMR data analysis

ERETIC correction ^21^ was applied to all DIRE spectra to ensure the observed intensities are quantitative. All DIRE spectra were calibrated by setting the spectral reference value to 0 (SR = 0). Integration of the *α* 1-acid glycoprotein N-acetyl signals GlycA and GlycB, and supramolecular phospholipid composite (SPC) was performed using scripts for the R Statistical Software ^22^ using standard functions. GlycA was cut at the following region *δ* 2.05*−*2.09, GlycB *δ* 2.09 *−* 2.12, SPC *δ* 3.15 *−* 3.35.

#### Biogenic amines, amino acids and tryptophan metabolic pathway analysis

Plasma samples were thawed at 4^*°*^C and prepared for analysis following previously reported methods for biogenic amines and amino acids ^23^ and tryptophan and associated catabolites ^24^. For the quantification of biogenic amines and amino acid metabolites, separation was performed by ultra-high-performance liquid chromatography (UHPLC) using an Acquity UPLC (Waters Corp., Milford, MA) coupled to a Bruker impact II QToF mass analyzer (Bruker, Daltonics, Billerica, MA). Resulting data files were processed for peak integration and quantification using Target Analysis for Screening Quantification (TASQ) software v2.2 (Bruker Daltonics, Bremen, Germany) where calibration curves were linearly fitted with a weighting factor of 1*/x*. For the measurement of tryptophan and associate catabolites, separation was performed using an Acquity UPLC (Waters Corp., Milford, MA) coupled to Waters Xevo TQ-XS mass spectrometer (Waters Corp., Wilmslow, U.K.). Obtained raw files were processed for peak integrations and metabolite quantification using TargetLynx package within MassLynx v4.2 (Waters Corp., Milford, MA) where calibration curves were linearly fitted using a weighting factor of 1*/x*. Resulting data matrices were combined and quality control checked prior to statistical analysis ^13,23^.

Three patients had at least a subset of their samples collected while being on parenteral nutrition (seven samples in total), which likely altered their lipid levels. Sensitivity analyses without these patients indicated that their exclusion leave our observations are unchanged.

### Data preprocessing and quality control

All statistical analyses were conducted using the R software ^22^. Except for ratios for which no systematic transformation was applied, CRP, cytokines and all other cellular and molecular variables were log-transformed for variance stabilization, using *x* ⟼ log_2_(*x* + 1) (with the offset “+1” accounting for zero counts while ensuring positivity). For each molecular dataset, the presence of extreme measurements and/or batch effects was assessed using principal component analysis (PCA) visualization. No batch effect was observed. A standard boxplot rule was applied to discard extreme samples, i.e., with *>* 20% of their measurements falling outside the Tukey outer fences. Following this procedure, 2 immune cell type samples (1.1%) and 10 metabolomic samples (1.6%) were removed from all downstream analyses (all glycoand lipoprotein samples were retained).

### Differential abundance analysis and association with CRP

Differential abundance analysis between COVID-positive patients and healthy controls was conducted using linear mixed modeling to account for serial subject measurements over a window of 7 weeks post symptom onset or positive swab. Analyses also included gender and age as fixed effects. The following model was implemented using `lmerTest` R package (R notation):

~~~
dep _ var ∼ covid _ status + age + gender + (1 | subject_id),
~~~

where the different molecular variables were taken in turn as the dependent variable `dep_var` and `covid_status` is a binary variable coding for “COVID positive” or “COVID negative”. Significance of the `covid_status` effect was assessed using a type 3 *F* test and Satterthwaite’s method (to estimate the degrees of freedom for fixed effects), and adjustment for multiple testing across each molecular dataset was performed using a false discovery rate (FDR) correction of 5%. When significance was reached, the molecular variable was called “upregulated” or “downregulated” based on the sign of the fold change.

The same linear mixed model framework was employed to test the association between CRP and each cellular/molecular variable, replacing the `covid_status` categorical variable with the quantitative (log-transformed) `CRP` variable.

Unless specified otherwise, all the analyses described hereafter are adjusted for multiple testing per data type, using an FDR correction of 5%.

### Functional principal component analysis

Functional principal component (FPC) analysis was conducted to characterize the inter- and intrapatient variability and estimate individual disease trajectories using parameters reflecting inflammation, as well as the immunometabolic response to infection. These parameters were CRP levels, five cytokines (IFN-*γ*, IL10, IL1B, IL6 and TNF), twelve lymphocyte subsets (CD4 EMRA, CD4 Naive, CD4 Non-naive HLA-DR^+^CD38^+^ T, CD8 EMRA, CD8 Naive, CD8 Non-naive HLA-DR^+^CD38^+^ T, CD19^+^, plasmablasts, gd T, MAIT, NK and NKT cells), three lipoproteins (HDA1, HDA2 and VLAB), two glycoprotein signals (GlycA and GlycB) and four metabolites from the kynurenine pathway (3-hydroxykynurenine, kynurenine, quinolinic acid and tryptophan).

The R packages `face`^25^ and `mfaces`^26^ were used to implement the univariate FPC analysis (for CRP levels), respectively multivariate FPC analysis (for all other groups of parameters listed above), thereby leveraging shared signals across molecular markers of a same type. Briefly, this analysis aimed at disentangling the main contributions to the data variability while estimating subject-level trajectories based on sparse observations. The method was unsupervised and made full use of the longitudinal measurements collected for each patient. The patient-level trajectory of each parameter was modeled as mean function plus a truncated sum of random deviations from the mean. These deviations were expressed as a linear combination of orthonormal eigenfunctions, weighted by patient-specific scores. The eigenfunctions accounted for the principal sources of variation in the data, and the scores conveyed how each patient’s trajectory deviated from the population mean. In the multivariate setting, each molecular parameter had a corresponding set of eigenfunctions but the scores were common to all parameters (and were not indexed by time).

In many situations including ours, the first FPC eigenfunctions capture nearly all of the variation. These eigenfunctions may also be interpretable; for all the above parameters, the major mode of variation pertained to disease severity (first eigenfunction and corresponding scores), while the second mode of variation reflected the type of “recovery profile” or recovery of the analyzed parameter(s) (second eigenfunction and corresponding scores). The FPC method also inferred smooth estimates of variance and auto-correlation functions. Cross-correlation functions could also be estimated in the multivariate case and provided insights on the covariation of different related molecular parameters over time. Asymptomatic patients (severity class A) were not considered for FPC analyses and all downstream analyses to avoid ambiguity when discussing recovery, and FPC trajectories were estimated over a time window of 7 weeks post symptom onset.

### Hierarchical clustering

Hierarchical clustering was performed on the CRP scores corresponding to the first two eigenfunctions to uncover groups of patients with similar disease trajectories. Complete linkage clustering was employed and the number of clusters was assessed using three diagnostics (Fig. S4): the gap statistics (suggested 3 clusters), the average silhouette width (suggested 2 clusters) and the total within sum of square (suggested 2 or 3 clusters). The three clusters corresponded to groups *i, ii* and *iii*, as defined in the Results section, and the two clusters corresponded to group *iii* and group *i+ii*, i.e., to merging the two groups with favorable evolution (the latter two clusters were employed for prediction modeling of systemic recovery).

Overall and one vs all Fisher’s exact tests were used to characterize the groups based on gender, and anova and one vs all *t*-test were used to assess differences in age. Unless otherwise specified, *p*-value-based significance was labelled as follows on plots: **** if *p <* 0.0001, *** if *p <* 0.001, ** if *p <* 0.01 and * if *p <* 0.05. For all subsequent analyses, significance labels are based on FDR-adjusted *p*-values.

### Correlation tests

Correlation between the parameters analyzed by FPC (lymphocytes, lipoproteins, glycoproteins, kynurenine-pathway metabolites, cytokines and CRP levels) was assessed during acute infection (0 *−* 21 days post symptom onset) and convalescence (22 *−* 49 days post symptom onset). For each time window and each subject, the multiple measurements per subject were averaged, prior to computing correlation within recovery groups. For healthy controls, samples were available at a single timepoint for each subject so a single correlation matrix was computed. FDR-adjusted correlation tests were performed using the R package `TestCor` separately for each recovery group and for the healthy controls.

Pairwise Pearson correlation tests among the severity and recovery FPC scores for lymphocytes, lipoproteins, glycoproteins, metabolites from the kynurenine pathway, as well as cytokines and CRP were also conducted.

### Longitudinal modeling

Univariate mixed models were employed to estimate the temporal profile of each cellular/molecular parameter for the recovery groups *i, ii* and *iii*, i.e., with random effects to account for serial measurements of patients. Polynomial splines of degree 2 were used to model the parameters with respect to the interaction between the time from symptom onset (`time`) and the recovery groups (`group`). The following model was fitted for each molecular variable (`dep_var`):

~~~
dep _ var ∼ time * group + (1 | subject_id).
~~~

The significance of baseline effects (i.e., difference between groups at time zero) and interaction effects (i.e., difference in the group temporal courses) were tested using likelihood ratio tests and adjusted for multiplicity across all variables of same data type.

Direct comparisons with healthy control levels were also conducted using mixed models whereby samples were grouped into five time windows (involving similar numbers of samples): (0, 3], (3, 7], (7, 12], (12, 27] and (27, 52] weeks post symptom onset. For each molecular variable and each time window, the following model was fitted:

~~~
dep _ var ∼ category + (1 | subject_id),
~~~

where `category` is a categorical variable coding for the healthy control and three recovery groups, with the former used a reference factor level. To assess the discrepancy between each group’s parameters and healthy control parameters, the significance of the `category` levels was examined, adjusting for multiplicity over all parameters of same data type. Significance was not reported if *≤* 15 samples were available in the group and the time window under consideration.

### Long-term symptoms

Long-term symptoms were collected under the form of questionnaires given to patients between 2 and 11 months post symptom onset (average: 6 months). Patients were asked to rank the severity of a list of symptoms on an ordinal scale. These scales were recoded so the scores range from 0 (no symptom) to 5 (extreme manifestation of the symptom) and a composite score was computed as the average of the individual symptom scores. The list of symptoms was: dyspnoea; cough; chest pain on exertion; palpitations or swollen ankles; persisting fever (2 months or more); new leg swelling in one leg or shortness of breath with chest pain; new skin rashes or sores; voice alteration; difficulties eating, drinking or swallowing; constant noisy breathing or throat whistling; anosmia or dysgeusia; difficulty to gain or maintain weight, loss of appetite; new neurology in one or more limbs; new pain in one or more parts of the body; muscle weakness, balance or range of movement of joints; fatigue; cognition: memory, concentration and thinking skills. Up to three questionnaires per patient were collected, but most patients completed a single questionnaire; the scores of patients with more than one questionnaire were averaged prior to the analysis. Non-parametric tests were used for overall comparison across recovery groups (Kruskall—Wallis rank sum test), as well as for pairwise tests (Wilcoxon rank sum tests).

### Survival analysis

The risk of death was studied by Kaplan—Meier survival analysis, using the three recovery groups *i, ii* and *iii* as strata. Difference in survival between the groups was assessed using a log-rank test.

### Predictive modeling

Predictive modeling of systemic recovery was carried out to classify patients, shortly after disease onset, in terms of unfavorable (group *iii*) or favorable disease progression (groups *i* and *ii*, merged). An integrative sparse generalized canonical correlation (sGCCA) approach, adapted for supervised analysis, DIABLO^27^, was applied on the first sample of each patient from the cohort, provided that this sample was taken within 3 weeks post symptom onset. Patients were randomly assigned to a training set or a left-out test set, according to a 70% *−* 30% split (using the R package caret to balance the recovery-category distributions within the training and test sets). sGCCA accounts jointly for the different data types (here: immune cell-types, polar metabolites, glyco- and lipoproteins, and selected metabolic ratios) and estimates integrative predictive signatures under the form of sparse latent components. The features composing these signatures might represent important molecular drivers of the type of recovery profile and their selection within a same signature might reflect shared underlying biological responses to infection, that simultaneously mobilize or affect features from the immune system, the metabolome and the lipidome.

Here the sGCCA method was trained to maximize the correlation between the different data types (cellular/molecular candidate predictors) as well as the discrimination between the recovery groups *i+ii* and *iii*. The training procedure used 3-fold cross-validation to select the numbers of candidate predictors within each latent component; the first two components were sufficient to achieve high discrimination between the recovery groups, and adding a third component didn’t yield further improvement.

### Systemic recovery prediction tool

The predictive model based on Cambridge patients’ early samples allows generating predictions for new samples collected when a patient presents to the clinic. An interactive tool to browse recovery prognoses is provided at http://shiny.mrc-bsu.cam.ac.uk/apps/covid-systemic-recovery-prediction-app. For a new patient, each marker from the two signatures identified by the model (Fig. 5A) can be set in terms of percentiles of the empirical distribution formed by all Cambridge cohort patient and healthy control measurements. The colors appearing on the bar suggest normal ranges (grey, corresponding to the healthy controls’ interquartile range), low values (blue, smaller than the healthy controls’ first quartile) or high values (red, larger than the healthy controls’ third quartile). The initial values correspond to the median of healthy controls’ measurements. The tool runs the prediction based on the input values and it outputs the systemic recovery prognosis (*i+ii* or *iii*), along with a predicted score ranging from 0.5 to 1 and conveying the degree of confidence about the prediction (the larger, the higher the confidence).

As only a subset of markers from the two signatures may be quantified from blood tests collected in the clinic, the prediction can be based on a selection of markers chosen from the drop-down menus; the deselected markers are omitted in the linear combination corresponding to their latent component. To assess the expected performance of the model when restricted to a subset of markers, ROC curves are recomputed based on this subset using the Cambridge left-out test set (the curves are updated when selecting or deselecting markers). A poor performance on the test set (e.g., AUC *<* 0.7) suggests that the selection of markers is insufficient to provide reliable predictions for the new patient. In that case, the predicted category should be disregarded and values for additional markers should be supplied where possible. As Vg9Vd2(hi) gd T appears in both signatures, its slider bar is displayed in the first column only.

## Data Availability

The data and metadata for the early (0-3 months) follow-up are available at NIHR CITIID COVID-19 Cohort (https://www.covid19cellatlas.org/patient/citiid/).

## Data availability

The data and metadata for the early (0-3 months) follow-up^9^ are available at NIHR CITIID COVID-19 Cohort (https://www.covid19cellatlas.org/patient/citiid/).

## Consortia

The members of the Cambridge Institute of Therapeutic Immunology and Infectious Disease-National Institute of Health Research (CITIID-NIHR) COVID BioResource Collaboration are John Allison, Ali Ansaripour, Stephen Baker, Laura Bergamaschi, Ariana Betancourt, Sze-How Bong, Georgie Bower, John R. Bradley, Ashlea Bucke, Ben Bullman, Katherine Bunclark, Helen Butcher, Jo Calder, Laura Canna, Daniela Caputo, Debbie Clapham-Riley, Chiara Cossetti, Jerome D. Coudert, Eckart M.D.D. De Bie, Aloka De Sa, Eleanor Dewhurst, Giovanni di Stefano, Jason Domingo, Gordon Dougan, Benjamin J. Dunmore, Anne Elmer, Madeline Epping, Codie Fahey, Stuart Fawke, Stewart Fuller, Anita Furlong, Nick Gleadall, Ian G Goodfellow, Stefan Graf, Barbara Graves, Jennifer Gray, Richard Grenfell, Ravindra K. Gupta, Julie Harris, Christoph Hess, Sarah Hewitt, Andrew Hinch, Josh Hodgson, Elaine Holmes, Christopher Huang, Oisín Huhn, Kelvin Hunter, Tasmin Ivers, Sarah Jackson, Isobel Jarvis, Emma Jones, Jane Kennet, Sherly Jose, Masa Josipovic, Mary Kasanicki, Nathalie Kingston,Jenny Kourampa, Elisa Laurenti, Ekaterina Legchenko, Paul J. Lehner, Emma Le Gresley, Daniel Lewis, Rachel Linger, Paul A. Lyons, Michael Mackay, John C. Marioni, Jimmy Marsden, Jennifer Martin, Cecilia Matara, Nicholas J. Matheson, Anne Meadows, Sarah Meloy, Nicole Mende, Federica Mescia, Alice Michael, Rachel Michel, Lucy Mwaura, Francesca Muldoon, Francesca Nice, Criona O’Brien, Ciara O’Donnell, Georgina Okecha, Ommar Omarjee, Nigel Ovington, Willem H. Owehand, Sofia Papadia, Caroline Patterson, Marianne Perera, Isabel Phelan, Linda Pointon, Petra Polgarova, Gary Polwarth, Nicole Pond, Jane Price, Cherry Publico, Rebecca Rastall, Carla Ribeiro, Nathan Richoz, Veronika Romashova, Sabrina Rossi, Jane Rowlands, Valentina Ruffolo, Caroline Saunders, Natalia Savinykh Yarkoni, Rahul Sharma, Joy Shih, Mayurun Selvan, Kenneth G.C. Smith, Sarah Spencer, Luca Stefanucci, Hannah Stark, Jonathan Stephens, Kathleen E Stirrups, Mateusz Strezlecki, Charlotte Summers, Rachel Sutcliffe, James E. D. Thaventhiran, Tobias Tilly, Zhen Tong, Hugo Tordesillas, Carmen Treacy, Mark Toshner, Paul Townsend, Lori Turner, Neil Walker, Jennifer Webster, Michael P. Weekes, Nicola K. Wilson, Jennifer Wood, Marta Wylot, and Cissy Yong.

## Acknowledgements

We thank all the patients and Health Care Workers who consented to take part in this study. We are grateful for the generous support of CVC Capital Partners, the Evelyn Trust (20/75), UKRI COVID Immunology Consortium, Addenbrooke’s Charitable Trust (12/20A), the NIHR Cambridge Biomedical Research Centre and the UKRI/NIHR through the UK Coronavirus Immunology Consortium (UK-CIC) for their financial support. H.R. is funded by the Lopez—Loreta Foundation. We would also like to thank the NIHR Cambridge Clinic Research Facility outreach team for enrolment of patients; and the NIHR Cambridge Biomedical Research Centre Cell Phenotyping Hub and the CRUK Cambridge Institute flow cytometry core facility for their support with flow and mass cytometry.

