## Supplementary Material for "A patient-centric characterization of systemic recovery from SARS-CoV-2 infection"

#### Figure S1

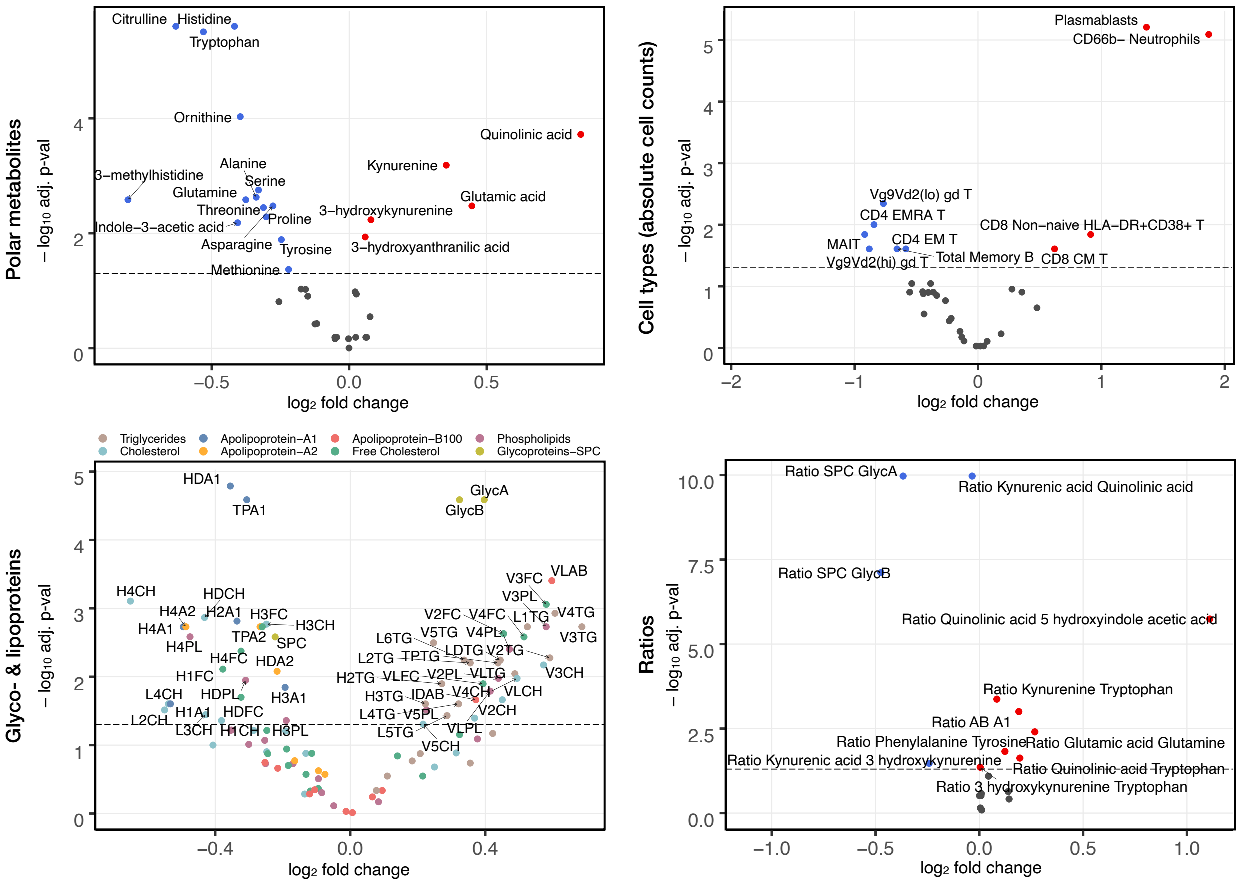


Figure S1: **Volcano plots for differentially abundant immune cell subsets, glyco- & lipoproteins, polar metabolites and diverse metabolic ratios, comparing COVID-positive patients with healthy controls.** The $x$-axis shows the $\log_2$ fold-change and the $y$-axis shows the $-\log_{10}$ adjusted $p$-value. The dashed horizontal line corresponds to a 5% FDR threshold, and red and blue indicate significant up- and downregulation in infected individuals, respectively. For lipoproteins, colors instead refer to compounds.

### Figure S2

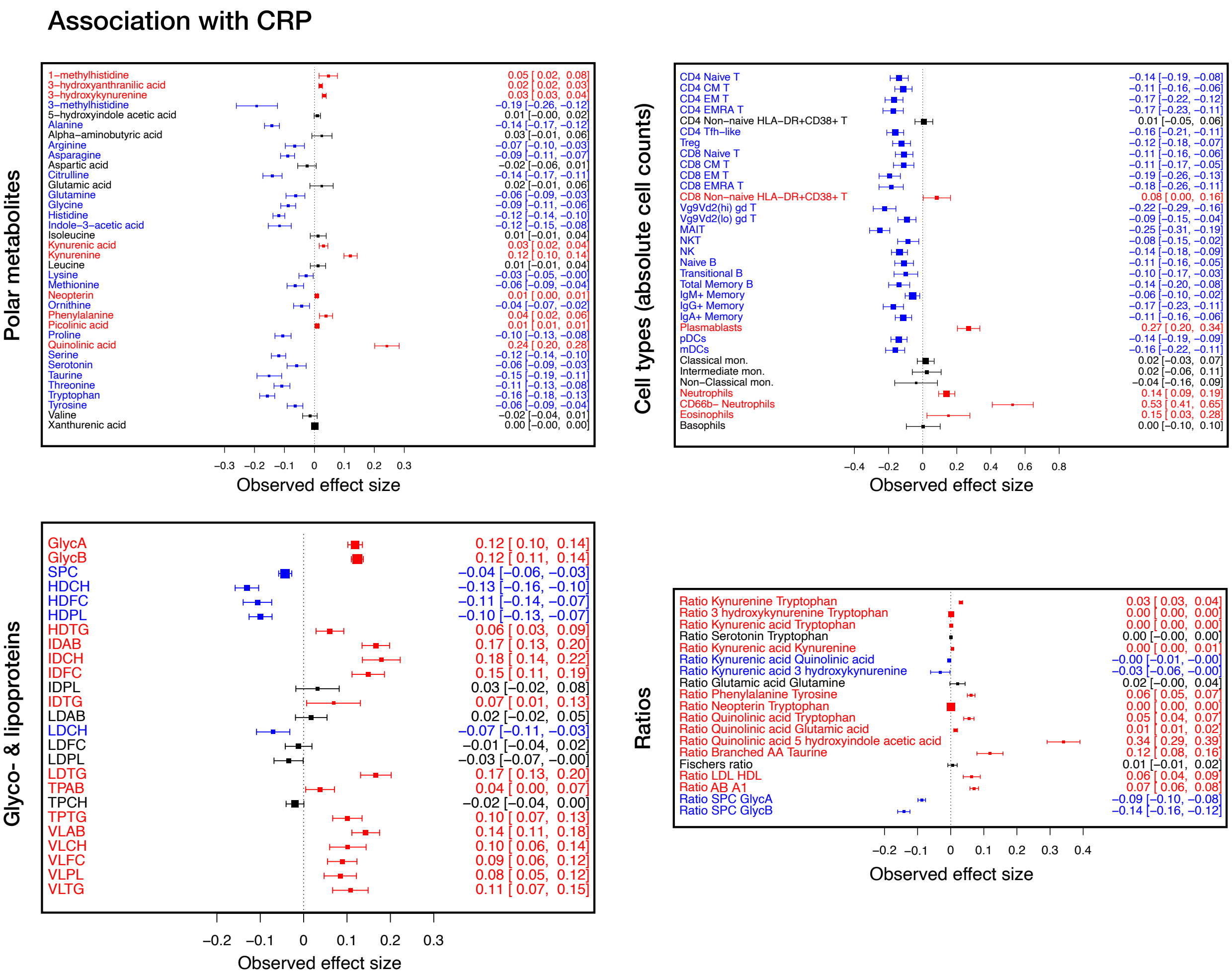


Figure S2: **Association of immune cell subsets, main classes of glyco- & lipoproteins, polar metabolites and diverse metabolic ratios with CRP abundance.** Red and blue indicate positive and negative effect, respectively, at FDR 5%.

#### Figure S3

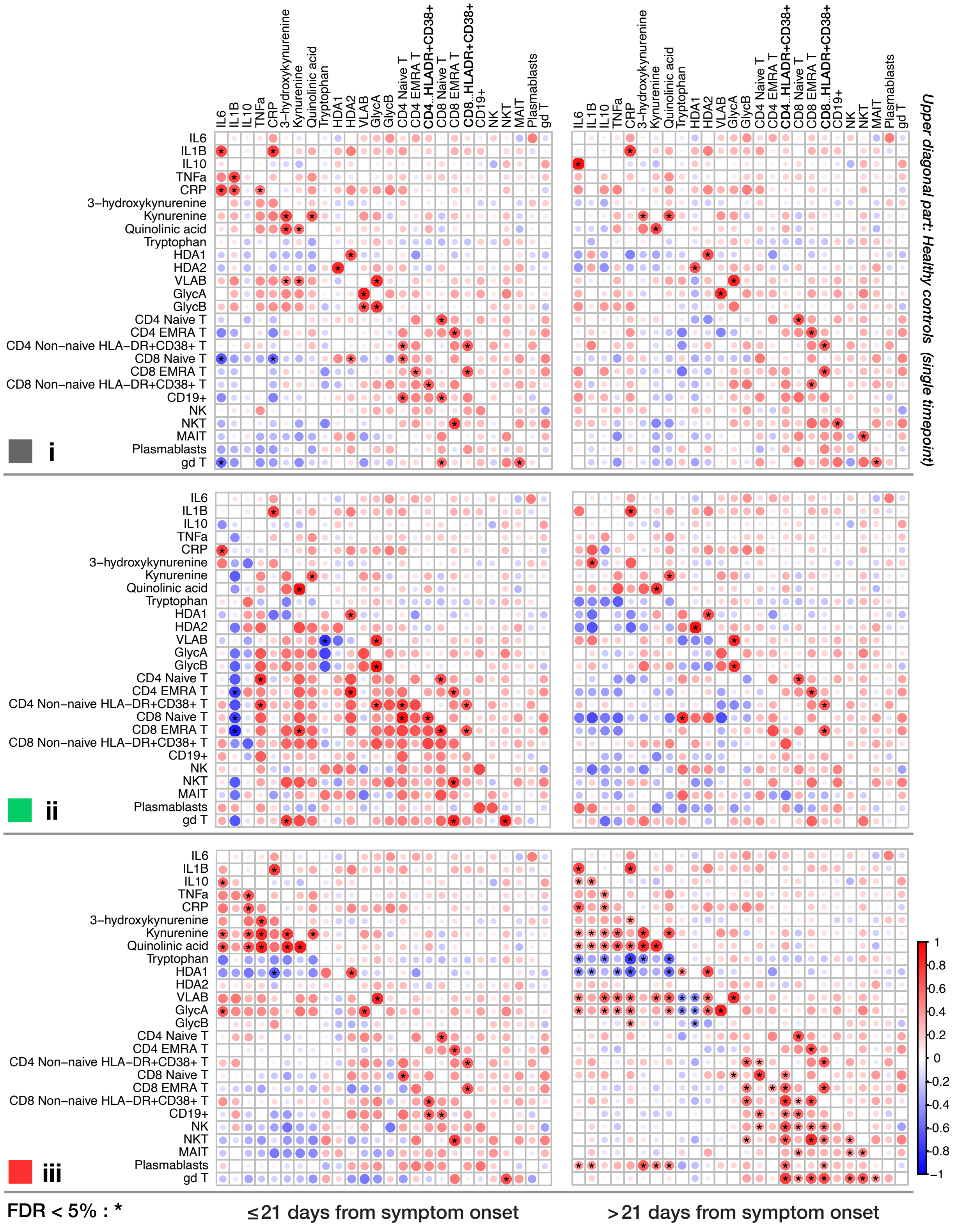


Figure S3: **Correlation among the variables modeled with the FPC approach for the three recovery groups and two time windows (0 − 3 weeks and 3 − 7 weeks post symptom onset).** The lower diagonal part of the matrices shows correlations from the patients' data while the upper diagonal part corresponds to correlations from the healthy controls' data and is the same for all matrices. The stars indicate significance at FDR 5%.

### Figure S4

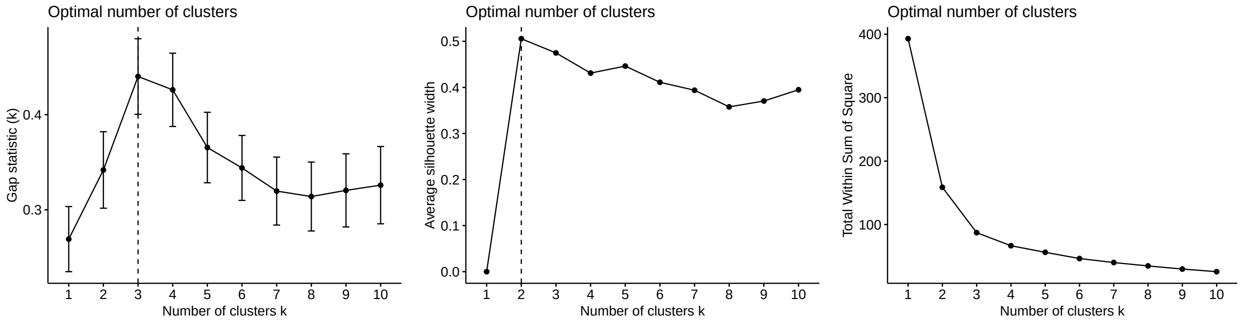


Figure S4: **Three diagnostics for the number of recovery groups obtained by hierarchical clustering the CRP FPC scores.** Gap statistic (suggests 3 clusters), the average silhouette width (suggests 2 clusters) and the total within sum of square (suggests 2 or 3 clusters). The three clusters correspond to the recovery groups *i*, *ii* & *iii*, and the two clusters corresponds to group *iii* (unfavorable disease progression) and merged group *i*+*ii* (favorable disease progression).
